# Impact of COVID-19 related maternal stress on fetal brain development: A Multimodal MRI study

**DOI:** 10.1101/2022.10.26.22281575

**Authors:** Vidya Rajagopalan, William T. Reynolds, Jeremy Zepeda, Jeraldine Lopez, Skorn Ponrartana, John Wood, Rafael Ceschin, Ashok Panigrahy

## Abstract

**Background:** Disruptions in perinatal care and support due to the COVID-19 pandemic was an unprecedented but significant stressor among pregnant women. Various neurostructural differences have been re-ported among fetuses and infants born during the pandemic compared to pre-pandemic counterparts. The relationship between maternal stress due to pandemic related disruptions and fetal brain is yet unexamined.

**Methods:** Pregnant participants with healthy pregnancies were prospectively recruited in 2020-2022 in the greater Los Angeles Area. Participants completed multiple self-report assessments for experiences of pandemic related disruptions, perceived stress, and coping behaviors and underwent fetal MRI. Maternal perceived stress exposures were correlated with quantitative multimodal MRI measures of fetal brain development using ltivariate models.

**Results:** Fetal brain stem volume increased with increased maternal perception of pandemic related stress positively correlated with normalized fetal brainstem volume (suggesting accelerated brainstem maturation). In contrast, increased maternal perception of pandemic related stress correlated with reduced global fetal brain temporal functional variance (suggesting reduced functional connectivity).

**Conclusions:** We report alterations in fetal brainstem structure and global functional fetal brain activity associated with increased maternal stress due to pandemic related disruptions, suggesting altered fetal programming. Long term follow-up studies are required to better understand the sequalae of these early multi-modal brain disruptions among infants born during the COVID-19 pandemic.

## 1. Introduction

The COVID-19 pandemic created many, unprecedented disruptions to everyday life particularly in 2020-2022 before vaccines were widespread. In addition to disruptions around employment, childcare, housing, and nutrition, pregnant women also suffered negative experiences related to support and care during pregnancy and childbirth. Social isolation, reduced access to child and elder care, COVID-19 infection risk, and changes to medical policies around pre and postpartum care were reported to be the most common stressors among pregnant women [1,2]. Pregnant women are particularly vulnerable to mood and anxiety related disorders [3] which are exacerbated during natural disasters or stressful events [4,5]. Unsurprisingly, pregnant women indicated elevated levels of stress during the COVID-19 pandemic [6]. In addition to health consequences for the mother, increased maternal stress has an intergenerational impact on fetal development [7,8]. Increased maternal stress during pregnancy is known to alter the fetal brain and adversely impact postnatal neurodevelopmental outcomes [9–12].

Studies of infants born during the COVID-19 pandemic have reported reduced cognitive, motor, and emotional development compared to those born pre-pandemic [7,8], with increased prenatal stress directly associated with adverse effect and temperament [13,14]. Simultaneously, changes to brain structure and function have also been reported in infants born during the pandemic [15]. Lu et al.[16] reported volumetric reductions in the brain among fetuses of women pregnant during the pandemic compared to a pre-pandemic cohort. Their findings showed a negative relationship between general ma-ternal stress and fetal brain volumes. However, their cohort did not show an increase in maternal stress or anxiety during a pandemic, and they did not measure maternal stress or anxiety specifically linked to the pandemic. Additionally, there is no data on if or how emerging functional networks in the fetal brain, which are known to be sensitive to ma-ternal stress, were impacted by pandemic related maternal stress. Early aberrations to functional organization of the brain are well known to have deleterious downstream ef-fects in brain and behavioral development. As such, a multimodal imaging study is im-portant to better understand how prenatal maternal stress sets up the offspring’s brain for a trajectory of compounding aberrant development.

Understanding the impact of pandemic related maternal stress on fetal development al-lows us to identify risk and resilience factors to mitigate maternal stress and consequently minimize the intergenerational effect of pandemic related stress. Coping behaviors, in response to stressful events, are known to be modifiable targets to mitigate maternal stress and anxiety [17,18]. Given the extraordinary nature of pandemic related stressors, there is little information on various coping behaviors that pregnant women have adopted during the pandemic [19–21]. Despite its observational nature, information on coping behaviors to pandemic related stressors allow clinical care teams to design and implement support programs aimed at improving maternal mental health during pregnancy and child out-comes.

In this work, we investigated the impact of maternal stress due to pandemic related dis-ruptions in pregnancy support and care on structural and functional development of the human fetal brain. Our primary hypothesis is that increased maternal stress would pre-dict quantitative alterations in structural and functional characteristics of the fetal brain. Secondarily, we compared coping behaviors between pregnant women reporting high vs low levels of pandemic related stress.

## 2. Materials and Methods

### 2.1 Subject Demographics

Pregnant mothers, living in the greater Los Angeles area were recruited using flyers, social media ads, and referrals from community partner clinics at Children’s Hospital Los Angeles (CHLA) from November 2020 – November 2021. Enrollment eligibility included healthy, pregnant women between 18 – 45 years with singleton, uncomplicated pregnancies (confirmed by ultrasound) between 21 – 38 gestational weeks (GW). Exclusion criteria were multiple gestation, fetal or genetic anomalies, congenital infection, and maternal contraindication to MRI. Informed consent for the study was obtained under a protocol approved by the Institutional Review Board at CHLA. Demographics, perinatal health history, and self-assessment surveys of consented participants were gathered via online survey within 24 hours prior to MRI.

### 2.2 Stress and Coping Behavioral Assessments

Participants were asked to complete the Coronavirus Perinatal Experiences - Impact Survey[22] (COPE-IS). This is a self-assessment questionnaire, available in multiple lan-guages, to assess feelings and experiences of pregnant women and new mothers in rela-tion to disruptions caused by the COVID-19 pandemic. Questions in this assessment were adapted from multiple validated questionnaires such as the Brief Symptom Inventory[23] PTSD checklist from DSM-5 [24], and the Johns Hopkins Mental Health Working Group. In this study, we only included questions pertinent to the prenatal period. Perceived maternal stress was computed as described here [21,22] and will be referred to as COPE-Stress going forward. Participants also completed the Brief COPE questionnaire[25], which is an abbreviated form of the COPE (Coping Orientation to Problems Exposed) questionnaire[26]. This is a self-assessment of a wide range of coping behaviors including both maladaptive coping (includes substance use, venting, behavioral disengagement, denial, self-blame, and self-distraction)[27] and adaptive coping (includes humor, planning and seeking social support, use of emotional and instrumental support, positive reframing, religion, and acceptance)[28,29]. This questionnaire has been validated in multiple languages and cultural contexts to be correlated to perceived stress and mental well-being.

### 2.3 Child Opportunity Index (COI)

Neighborhood socio-economic environment (SEE) is a known modifier of overall maternal stress during pregnancy[30], pandemic related stress[31], and offspring out-comes[32]. Family income is often used to measure SEE. However, the quality of life associated with absolute income number varies regionally based on cost of living, social policies, environmental factors, etc. To overcome these limitations, we chose to represent SEE using childhood opportunity index (COI). COI is a multi-dimensional, nationally normed measure of the quality of social, environmental, health, and educational resources available at each zip code[33]. We extracted maternal COI using self-reported zip code at the time of the MRI visit and will be referred to as COI-SEE going forward.

### 2.4 Image Acquistion

Pregnant mothers were prospectively recruited between 24-38 GW and imaged on 3.0 T Philips Achieva scanner (Netherlands). Multiplanar single-shot turbo spin echo imaging was per-formed (TE = 160 ms, TR = 9000-12,000 ms, 3 mm slice thickness, no interslice gap, 1 × 1 mm in plane resolution). Fetal brains were scanned in each of three planes for three times resulting in nine images per subject and images were repeated if excessive motion was present. Echo-planar imaging (EPI) BOLD images were also collected with the following parameters: FOV = 300mm TR = 2000 ms, TE = 31-35 ms (set to shortest), flip angle = 80o, with an in-plane resolution of 3×3 mm2, slice thickness of 3.0 mm and 0.0 mm intra-slice gap. 150 timepoints were recorded for each BOLD image and two images were collected for each subject.

### 2.5 Image Processing

#### 2.5.1 Brain Structure

All structural brain images were verified as being typical for gestational age by a board certified neuroradiologist (SP). For each subject, various 2D stacks of the T2 images were visually assessed to identify and discard stacks with large, spontaneous fetal motion. In each stack, the fetal brain was localized from surrounding tissue. For each subject, multiple 2D stacks were motion corrected and reconstructed, using a slice-to-volume reconstruction [34] into a 3D volumetric T2 image with an isotropic resolution of 1 mm^3^. Reconstructed fetal brains were processed through a bespoke, automated fetal segmentation pipeline. Each fetal brain was normalized (affine followed by non-rigid) to a probabilistic atlas [35] of equivalent gestational age using Advanced Normalization tools[36]. Segmentations were manually inspected for accuracy and subjects with failed segmentations were discarded. The resulting segmentation maps were subsequently refined. To ensure consistency across different gestational ages, transient structures only present in the tissue atlas from 21 – 30 weeks of gestation such as the subplate, intermediate zone, and ventricular zone were combined with the corpus collosum and labeled as developing WM (WM). Cerebrospinal fluid (CSF) segmentation was refined as intra-ventricular (within lateral ventricles) and extra-axial CSF. Due to the small size and relative difficulty in segmenting the hippocampus and amygdala, both structures were combined into a hippocampus-amygdala complex. Deep grey tissue was defined as the combination of the caudate, putamen, thalamus, fornix, internal capsule, subthalamic nucleus, and hippocampal commissure. Right and left hemispheric labels were combined into a single volume for each structure. The final segmentation yielded volumes of the following structures: cortical plate, developing white matter, intra-ventricular CSF, extra-axial CSF, deep gray tis-sues, cerebellum, hippocampal-amygdala complex, and brainstem. A total brain volume (TBV) was generated for each subject as the sum of all tissues.

#### 2.5.2 Brain Function

BOLD imaging of the fetal brain is prone to spontaneous fetal motion which is com-pounded by lower signal to noise ratio and spatial resolution. While modern motion cor-rection algorithms effectively attenuate the effects of subject motion on the temporal data, they are limited in effect beyond small degrees of motion. Any robust voxel-wise approach to functional fetal imaging would yield a prohibitively low number of subjects with usable data. We therefore chose to implement a whole-brain temporal signal approach to fetal functional imaging. Resting state images were first motion corrected using FSL’s MCFLIRT routine, using the first frame as the registration target, and a mean framewise displacement threshold > 0.2 mm to eliminate frames with excessive motion. As the intent of this study was to use minimally processed data using framewise measures, as opposed to voxelwise measures, we made no prior assumptions on physiological or nuisance frequency thresholds in fetal functional imaging, and did not apply any bandpass filtering. A mean brain signal image was then generated by averaging across every frame in the sequence. This mean signal image was used as the source image for brain extraction to generate a brain mask. Brain extraction was done by using an adaptive routine that iterated between using FSL’s Brain Extraction Tool (BET)[37] and AFNI’s Skullstrip, using decreasingly smaller thresholds for brain tissue [38]. This approach yielded a good approximation of the fetal brain, with a minimal manual correction step required for final brain masking. The brain mask was then propagated across each frame in the temporal sequence to extract only fetal brain voxels.

Using the mask generated above, we averaged the whole brain BOLD signal in each frame and generated statistical measures across time. The measures generated were temporal mean (average of the mean signal across frames), temporal variability (average of the standard deviation of the signal across frames), variance of the mean (variance of the mean signal in each frame), kurtosis of the mean (kurtosis of the mean signal in each frame). Finally, to test for any signal or physiological drift, we calculated the autocorrelation of the mean signal in each frame, and the kurtosis and autocorrelation of the normalized signal across frames.

### 2.6 Statistical Analysis

#### 2.6.1 Brain Structure

Regression analysis was performed in Python (3.7) using the Statsmodel.api v0.13.2. We used multiple, linear regression to model the relationship of COPE-Stress Score, COI-SEE, and their interaction on TBV after adjusting for gestational age at MRI. Nested models of the covariates without interaction were also tested. Models were deemed to be significant if one or more of the covariates were statistically significant, and models in-cluding the interaction term were only selected over the simpler counterpart if they had a higher explained variance (R-squared) and/or lower Bayes’ Information Criteria (BIC). Using similar regression models, we individually tested the relationship of COPE-Stress score and COI-SEE for each tissue volume listed in Section 2.4.1 (as a dependent variable). Secondarily, we also tested the relationship of COPE-Stress score and COI-SEE on tissue volumes normalized by TBV after adjusting for gestational age.

#### 2.6.2 Brain Function

Statistical analysis for brain functional metrics was similar to Section 2.5.1. A separate regression model was tested for each, individual functional metric (Section 2.4.2) with COPE Stress, COI-SEE, and their interaction as predictor variables after accounting for GA at MRI.

#### 2.6.3 Comparison of Coping Behaviors

Coping behaviors, both the Brief-COPE and COVID specific, were analyzed for differences between low and high stress mothers. Mothers were split into low, medium, and high stress categories based on tertiles of COVID Stress scores. Using Fischer Exact test, we compared if mothers reporting low and high stress used each coping behavior at significantly different amounts.

## 3. Results

### 3.1 Subject Demographics

Pregnant mothers were recruited prospectively for this study with a total of 45 mother-fetal dyads completed the MR imaging session. Three subjects had missing zip code information, and which resulted in missing COI-SEE data and was thus excluded from any analysis. After imaging, three subjects failed brain segmentation resulting in 39 sub-jects for structural regression results. A total of 43 subjects of the original 45 subjects had analyzable BOLD imaging and were used for the functional regression results (Table 1).

**Table 1.** Study participant demographics including maternal parity and maternal race/ethnicity.

| Characteristic | Total |  |
| --- | --- | --- |
| **Total Participants** | 45 |  |
| **Sex of fetus** |  |  |
| Female | 18 |  |
| Male | 20 |  |
| Unknown | 7 |  |
| **Total MRIs** | 45 |  |
| **GA, median (range), wk** |  |  |
| At MRI | 31.57 | (22.57 to 38.42) |
| At Birth | 39.14 | (33 to 41.86) |
| **Maternal age at MRI, median, yr** | 32 | (18 to 43) |
| **Maternal parity** |  |  |
| Primiparous | 18 |  |
| Multiparous | 22 |  |
| Unknown | 5 |  |
| **Infant Weight, median, kg** | 3.54 |  |
| **Mother's race/ethnicity** |  |  |
| Caucasian | 8 |  |
| Hispanic or Latino | 28 |  |
| Asian/Pacific Islander | 7 |  |
| African American | 1 |  |
| Middle Eastern | 0 |  |
| Other or unknown | 1 |  |

### 3.2 Brain Structure

There were no significant associations between absolute volumes of the various brain structures and perceived maternal stress, COI-SEE, or their interaction (Table 2). However, there was a significant positive association between normalized brain stem volume and perceived maternal stress (p = 0.03) but not with COI-SEE and the interaction of COI-SEE and maternal stress (Table 3) There were no significant associations between normalized volumes of other structures with COPE-Stress or COI-SEE.

**Table 2.** Raw brain structure volumes relationship to COVID stress and COI-SEE

| Volume (cm3) | COVID Stress Score |  | COI Nationally Normed Value |  | COI Stress Interaction |  |
| --- | --- | --- | --- | --- | --- | --- |
| | $\beta$ (CI) | P-Value | $\beta$ (CI) | P-Value | $\beta$ (CI) | P-Value |
| Brainstem | 3.89E+00,<br>(-7.62E+01,<br>8.40E+01) | 0.97 | -2.81E-01,<br>(-1.41E+01,<br>1.35E+01) | 0.99 | 4.06E-01, (-1.18E+00,<br>1.99E+00) | 0.86 |
| Cerebellum | 1.54E+02,<br>(-4.84E+01,<br>3.56E+02) | 0.61 | 3.28E+01, (4.13E-01,<br>6.52E+01) | 0.49 | -1.95E+00,<br>(-5.79E+00,<br>1.89E+00) | 0.73 |
| Cortical Plate | -7.33E+02,<br>(-1.35E+03,<br>-1.18E+02) | 0.42 | -3.78E+00,<br>(-1.59E+02,<br>1.52E+02) | 0.99 | 1.23E+01,<br>(-1.43E+00,<br>2.60E+01) | 0.55 |
| Deep Grey | 1.93E+01,<br>(-1.83E+02,<br>2.22E+02) | 0.95 | 2.76E+00,<br>(-3.17E+01,<br>3.73E+01) | 0.96 | 1.65E+00,<br>(-2.28E+00,<br>5.58E+00) | 0.78 |
| Extra Axial CSF | -7.29E+02,<br>(-1.74E+03,<br>2.81E+02) | 0.63 | -9.28E+01,<br>(-3.06E+02,<br>1.21E+02) | 0.77 | 1.76E+01,<br>(-5.28E+00,<br>4.04E+01) | 0.60 |
| Hippocampus amygdala complex | -1.31E+00,<br>(-2.72E+01,<br>2.46E+01) | 0.97 | -7.62E-01,<br>(-6.21E+00,<br>4.69E+00) | 0.92 | 2.17E-01, (-3.33E-01,<br>7.68E-01) | 0.79 |
| Intra ventricular CSF | 2.59E+01,<br>(-7.98E+01,<br>1.32E+02) | 0.87 | 1.23E+01,<br>(-1.15E+01,<br>3.60E+01) | 0.73 | -5.07E-01,<br>(-2.81E+00,<br>1.79E+00) | 0.88 |
| White Matter | -5.17E+02,<br>(-1.69E+03,<br>6.58E+02) | 0.77 | -8.47E+01,<br>(-2.99E+02,<br>1.30E+02) | 0.79 | 1.19E+01,<br>(-1.25E+01,<br>3.63E+01) | 0.74 |
| Total Brain Volume | -2.51E+03,<br>(-6.81E+03,<br>1.80E+03) | 0.69 | -2.27E+02,<br>(-1.05E+03,<br>5.96E+02) | 0.85 | 5.92E+01,<br>(-3.03E+01,<br>1.49E+02) | 0.66 |

**Table 3.**
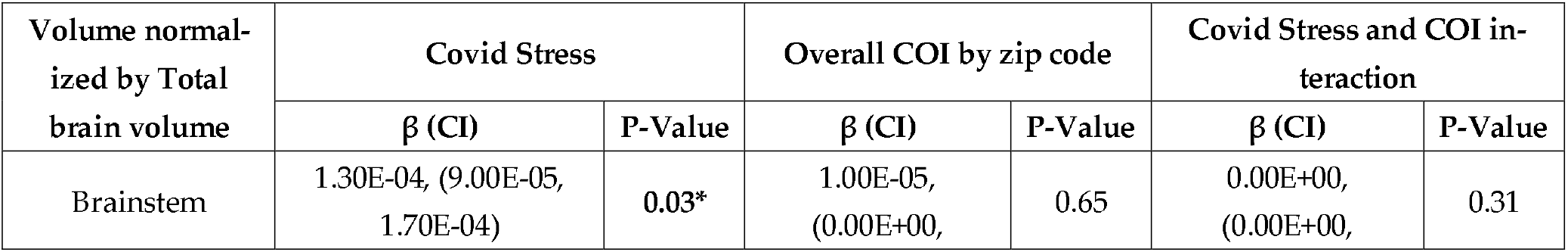
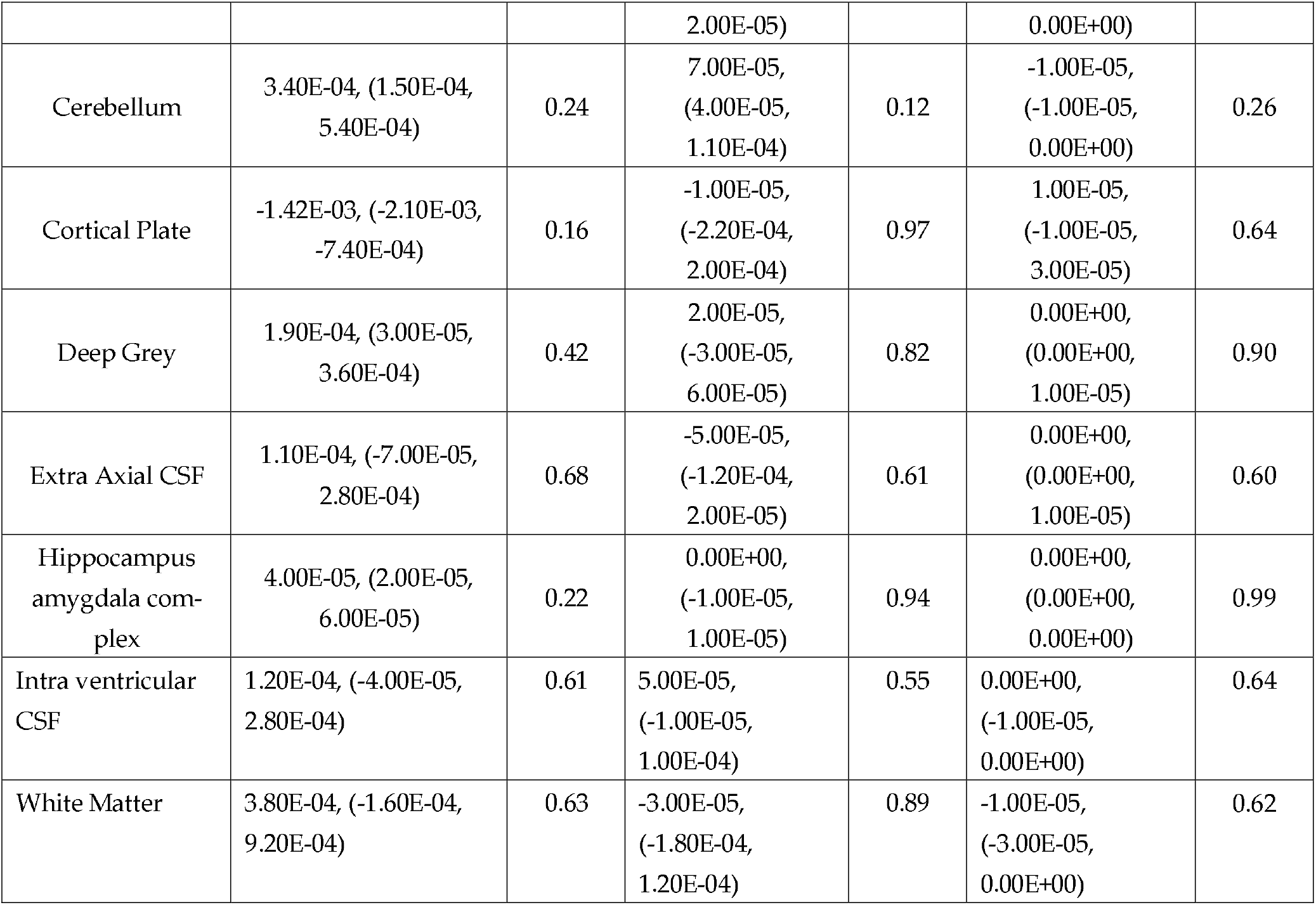
Brain structure volumes’, after normalization to total brain volume, relationship to COVID stress, COI-SEE, and their interaction

### 3.3 Brain Function

Lack of significant relationship between autocorrelation metrics and the predictor variable confirmed the absence of any systematic signal or physiological drifts. We found a significant negative relationship between temporal variability and COPE Stress (p < 0.028) (Table 4). The temporal variability model including the interaction term between Cope Stress Score and COI SES had a slightly improved R-squared (0.267) but lower BIC and reduced statistical significance of the covariates, likely due to co-linearity. We there-fore report the original model without the interaction term. We found no other statistically significant relationships between fetal brain functional characteristics with COPE Stress or COI SEE.

**Table 4.** Brain functional metrics’ relationship to COVID stress and COI-SEE using linear modeling.

|  | Covid Stress |  | Overall COI by zip code |  |
| --- | --- | --- | --- | --- |
| | $\beta$ (CI) | P-Value | $\beta$ (CI) | P-Value |
| Temporal mean of BOLD Signal | 135.369, (-509.52, 38.1) | 0.09 | 316.9634, (-604.97, 1238.9) | 0.49 |
| Temporal variability of BOLD Signal | -113.94, (-215.18, -12.71) | **0.03\*** | -19.5173, (-360.388, 321.354) | 0.91 |
| Variance of framewise mean BOLD signal | -5336.81, (-2.87e+04, 1.81e+04) | 0.65 | -5191.57, (-8.4e+04, 7.36e+04) | 0.9 |
| Kurtosis of framewise mean BOLD signal | 0.329, (-0.144, 0.802) | 0.17 | 0.457, (-1.135, 2.049) | 0.57 |
| Autocorrelation of framewise mean BOLD | -6.828e+06, (-1.41e+07, 4.89e+05) | 0.07 | 1.005e+07, (-1.46e+07, 3.47e+07) | 0.41 |

### 3.4 Comparison of Coping Behaviors

We compared coping behaviors between participants reporting high and low stress in our cohort. Among general coping behaviors measured by Brief-COPE, humor (p-value = 0.025) and venting (p-value = 0.048) were used more commonly by participants report-ing low stress compared to those reporting high stress (Figure 1). Among COVID specific coping behaviors that showed access to a mental health provider (p-value = 0.038), and information about how to reduce stress (p-value = 0.038) were chosen as being ‘Very Important’ to women reporting low stress at a high amount than in women reporting high stress (Figure 2). No other behaviors were found to be significantly different between high and low stress mothers. A full summary of the results can be seen in Figures 1 and 2.

**Figure 1.**
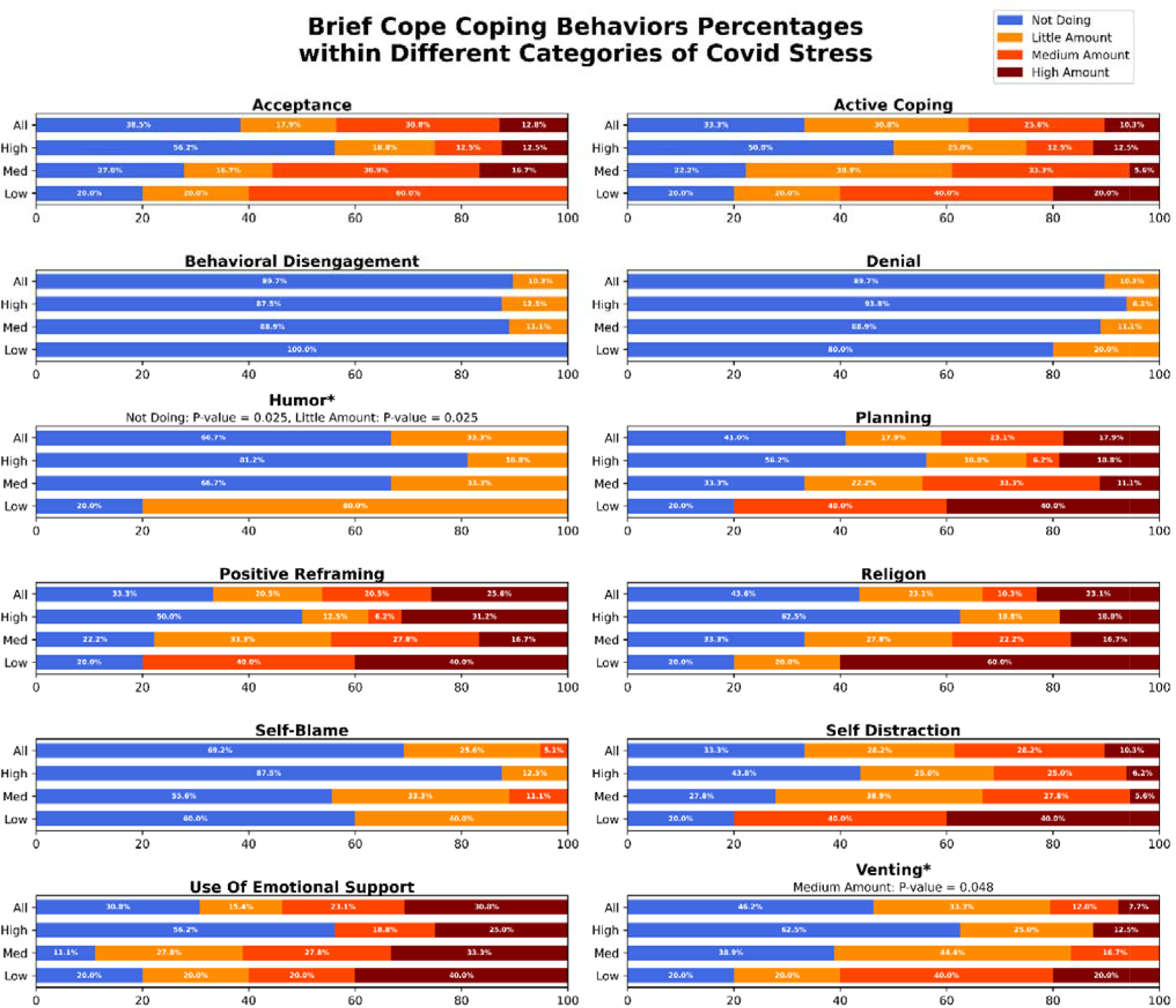
Comparison of general coping behaviors grouped by usage and analyzed for differences in incidence using a Fischer Exact Test.

**Figure 2.**
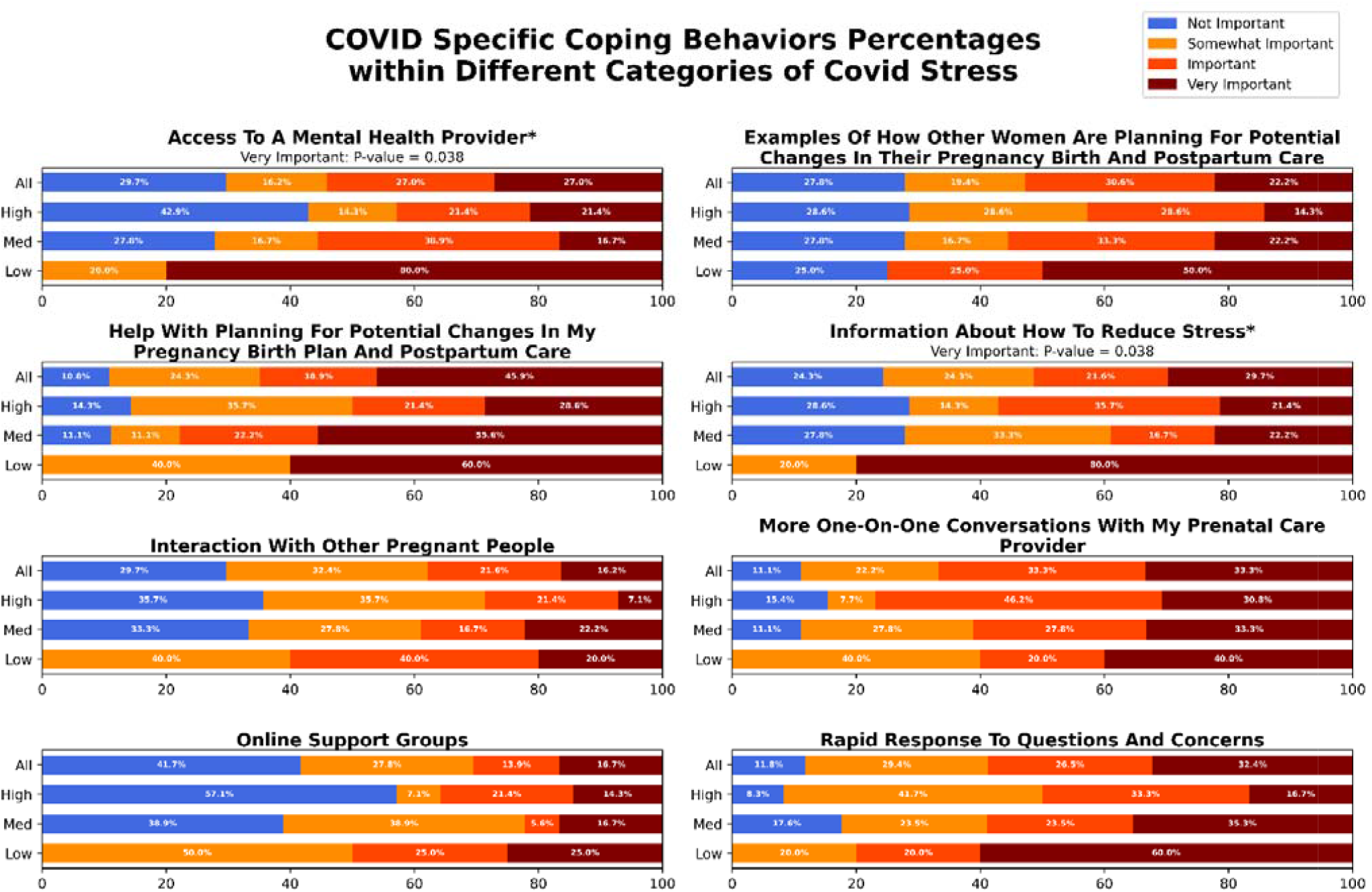
Analysis of COVID specific coping behaviors grouped by usage and analyzed for differences in incidence using a Fischer Exact Test.

## 4. Discussion

Our findings show that perceived maternal stress, in the setting of COVID-19 related care disruptions, impacts with structural and functional developmental of the fetal brain. Higher maternal stress was associated with increased brainstem volume (suggesting accelerated brainstem maturation) and globally decreased temporal variability of function (suggesting reduced functional connectivity) in the fetal brain. Additionally, we also found differences in the prevalence of specific coping behaviors between pregnant women who reported high stress compared to those who reported low stress.

We found that increased levels of maternal stress correlated with increased normalized brainstem volume suggesting relatively increased acceleration of brainstem maturation relative to cortical/supratentorial cerebral regions. Importantly, these results are con-sistent with prior studies that have correlated prenatal maternal stress and neonatal brainstem auditory evoked potentials (the speed at which the brainstem auditory evoked potential is conducted through the auditory nerve serves as a proxy for greater neural maturation)[39,40]. These studies have found significant relations between higher maternal prenatal distress and faster conductance, suggesting that greater maternal prenatal stress is associated with accelerated subcortical/brainstem neural maturation in neonates [41]. Our results are also consistent with the recent study by De Asis-Cruz et al. [42] which found that altered functional connectivity between brainstem and sensorimotor regionals were associated with high maternal anxiety scores.

We found that higher perceived maternal stress was associated with lower temporal variability in the fetal brain suggesting aberrations to foundational characteristics of con-nectivity and organization of emerging brain networks[43]. It has been well-established that such perturbations to early brain connectivity architecture, during the critical fetal period, has long-standing effects on behavioral and psychiatric development among these children[44–46]. Our findings of altered brain connectivity agree with previous findings of altered brain connectivity in infants of mothers who reported higher stress during the pandemic[15]. Behavioral and functional deficits particularly in the motor, cognitive and temperamental domain have been widely reported in various studies investigating the impact of maternal stress during the pandemic on child outcomes [7,8,13,14]. Increased maternal stress and anxiety traits (outside the setting of the pandemic) have been shown to alter functional architecture of the fetal brain[47]. Collectively, our and prior findings suggest that in utero alterations to brain architecture, associated with maternal stress during the pandemic, could underlie developmental deficits reported in these children. Further meta studies are needed to investigate the trajectory of brain development in children conceived and born during the pandemic.

Our findings suggest key differences in coping behaviors between pregnant women who reported low and high stress. Increased use of adaptive coping behaviors (particularly humor and venting) was more common among pregnant women who reported lower stress compared to those who reported higher stress. This association between in-creased use of adaptive, active coping and lower stress perception was reported across multiple studies of mental health in peripartum women during COVID-19 pandemic [21,48,49]. Our findings are also in agreement with generalized findings of positive rela-tionship between active coping behaviors and improved mental well-being in pregnant women[50]. In questions regarding COVID-19 specific coping behaviors, pregnant moth-ers reporting low stress endorsed access to mental health information and providers as being key to wellness. Routine screening for prenatal stress, provision of stress manage-ment information, and improved access to prenatal mental health care provide potential avenues for improving mental health and associated outcomes in pregnant women re-gardless of pandemic conditions.

This study’s limitations include small sample size and recruitment limited to a single geographical area in the USA during the pandemic. Since the greater Los Angeles area was disproportionately affected by pandemic related disruptions, comparison to a multi-site cohort will provide greater statistical power thereby increasing the generalizability of our findings. The cross-sectional nature of prenatal stress assessment limits our ability to associate time-varying stress levels and fetal outcomes. But all participating women became pregnant after pandemic-related restrictions were put in place. Lack of a pre-pandemic cohort limits our ability to pin-point if the differences in coping behaviors between pregnant women reporting low and high stress are specific adaptations to stress experienced during the pandemic.

## 5. Conclusions

Here, we reported the first multi-modal study of the impact of COVID-19 pandemic related maternal stress on fetal brain development. Our findings showed that increased maternal stress due to pandemic related disruptions was associated with structural and functional disruptions to fetal brain development and is suggestive of altered fetal pro-gramming. Comparing coping behaviors between pregnant women reporting higher and lower stress, our study provides insight into potential avenues for improved stress management and mental health outcomes among pregnant women.

## 6. Patents

This section is not mandatory but may be added if there are patents resulting from the work reported in this manuscript.

## Data Availability

All data produced in the present study are available upon reasonable request to the author

## Author Contributions

Conceptualization, VR, RC, and WR; methodology, VR, RC, and WR; validation, RC, SP, and WR.; resources, VR; data curation, VR, JZ, and JL; writing—original draft preparation, VR, RC, WR, and JZ; writing—review and editing, VR, WR, JZ, JW, RC, SP, JW and AP; visualization, VR, RC, WR and JZ.; supervision, AP and VR; funding acquisition, VR, AP. All authors have read and agreed to the published version of the manuscript.

## Funding

This work was funded NIH/NHLBI K01HL153942 and the Saban Research Institute Research Career Development Award to VR. Funding for WR was provided by the National Library of Medicine T15 Training program grant; award number 2T15LM007059-36. AP reported receiving grants from the Department of Defense (W81XWH-16-1-0613), the National Heart, Lung and Blood Institute (R01 HL152740-1, R01 HL128818-05), and Additional Ventures.

Institutional Review Board Statement: The study was conducted in accordance with the Declaration of Helsinki and approved by the Institutional Review Board (or Ethics Committee) of Childrens Hospital Los Angeles (protocol code CHLA-17-00292 and 9/14/2017).

## Informed Consent Statement

Any research article describing a study involving humans Written informed consent to include deidentified data has been obtained from the patient(s) to publish this paper

## Data Availability Statement

Due to limitations of informed consent, data from the study cannot be shared. But Methodologies and techniques from the study will be made available via direct email to the corresponding author.

## Acknowledgments

We would like to acknowledge all the women and families who participated in the study during an unprecedented global pandemic. We would like to thank Rosa Rangel, Teddy Aguilar, and Veronica Gonzalez for their help with the study.

## References

1. Zhou, J.; Havens, K.L.; Starnes, C.P.; Pickering, T.A.; Brito, N.H.; Hendrix, C.L.; Thomason, M.E.; Vatalaro, T.C.; Smith, B.A. Changes in Social Support of Pregnant and Postnatal Mothers during the COVID-19 Pandemic. Mid-wifery 2021, 103, 103162, doi:10.1016/J.MIDW.2021.103162.

2. Barbosa-Leiker, C.; Smith, C.L.; Crespi, E.J.; Brooks, O.; Burduli, E.; Ranjo, S.; Carty, C.L.; Hebert, L.E.; Waters, S.F.; Gartstein, M.A. Stressors, Coping, and Resources Needed during the COVID-19 Pandemic in a Sample of Pe-ri-natal Women. BMC Pregnancy Childbirth 2021, 21, 1–13, doi:10.1186/S12884-021-03665-0/TABLES/4.

3. Shorey, S.; Chee, C.Y.I.; Ng, E.D.; Chan, Y.H.; Tam, W.W.S.; Chong, Y.S. Prevalence and Incidence of Postpartum Depression among Healthy Mothers: A Systematic Review and Meta-Analysis. J Psychiatr Res 2018, 104, 235–248, doi:10.1016/J.JPSYCHIRES.2018.08.001.

4. King, S.; Dancause, K.; Turcotte-Tremblay, A.M.; Veru, F.; Laplante, D.P. Using Natural Disasters to Study the Ef-fects of Prenatal Maternal Stress on Child Health and Development. Birth Defects Res C Embryo Today 2012, 96, 273–288, doi:10.1002/BDRC.21026.

5. Salm Ward, T.; Kanu, F.A.; Robb, S.W. Prevalence of Stressful Life Events during Pregnancy and Its Association with Postpartum Depressive Symptoms. Arch Womens Ment Health 2017, 20, 161–171, doi:10.1007/S00737-016-0689-2.

6. Motrico, E.; Domínguez-Salas, S.; Rodríguez-Domínguez, C.; Gómez-Gómez, I.; Rodríguez-Muñoz, M.F.; Gómez-Baya, D. The Impact of the COVID-19 Pandemic on Perinatal Depression and Anxiety: A Large Cross-Sectional Study in Spain. Psicothema 2022, 34, 200–208, doi:10.7334/PSICOTHEMA2021.380.

7. Bianco, C.; Sania, A.; Kyle, M.H.; Beebe, B.; Barbosa, J.; Bence, M.; Coskun, L.; Fields, A.; Firestein, M.R.; Goldman, S.; et al. Pandemic beyond the Virus: Maternal COVID-Related Postnatal Stress Is Associated with Infant Tem-perament. Pediatric Research 2022 2022, 1–7, doi:10.1038/s41390-022-02071-2.

8. Shuffrey, L.C.; Firestein, M.R.; Kyle, M.H.; Fields, A.; Alcántara, C.; Amso, D.; Austin, J.; Bain, J.M.; Barbosa, J.; Bence, M.; et al. Association of Birth During the COVID-19 Pandemic With Neurodevelopmental Status at 6 Months in Infants With and Without In Utero Exposure to Maternal SARS-CoV-2 Infection. JAMA Pediatr 2022, 176, e215563–e215563, doi:10.1001/JAMAPEDIATRICS.2021.5563.

9. Schuurmans, C.; Kurrasch, D.M. Neurodevelopmental Consequences of Maternal Distress: What Do We Really Know? Clin Genet 2013, 83, 108–117, doi:10.1111/CGE.12049.

10. Talge, N.M.; Neal, C.; Glover, V. Antenatal Maternal Stress and Long-Term Effects on Child Neurodevelopment: How and Why? J Child Psychol Psychiatry 2007, 48, 245–261, doi:10.1111/J.1469-7610.2006.01714.X.

11. Wu, Y.; Lu, Y.C.; Jacobs, M.; Pradhan, S.; Kapse, K.; Zhao, L.; Niforatos-Andescavage, N.; Vezina, G.; du Plessis, A.J.; Limperopoulos, C. Association of Prenatal Maternal Psychological Distress With Fetal Brain Growth, Me-tabolism, and Cortical Maturation. JAMA Netw Open 2020, 3, e1919940–e1919940, doi:10.1001/JAMANETWORKOPEN.2019.19940.

12. van den Heuvel, M.I.; Hect, J.L.; Smarr, B.L.; Qawasmeh, T.; Kriegsfeld, L.J.; Barcelona, J.; Hijazi, K.E.; Thomason, M.E. Maternal Stress during Pregnancy Alters Fetal Cortico-Cerebellar Connectivity in Utero and Increases Child Sleep Problems after Birth. Scientific Reports 2021 11:1 2021, 11, 1–12, doi:10.1038/s41598-021-81681-y.

13. Griffin, M.; Ghassabian, A.; Majbri, A.; Brubaker, S.G.; Thomason, M. Evaluating the Association Between Ma-ternal Peripartum Care Interruptions and Infant Affect: A Longitudinal Study. Am J Obstet Gynecol 2022, 226, S310, doi:10.1016/J.AJOG.2021.11.521.

14. Provenzi, L.; Grumi, S.; Altieri, L.; Bensi, G.; Bertazzoli, E.; Biasucci, G.; Cavallini, A.; Decembrino, L.; Falcone, R.; Freddi, A.; et al. Prenatal Maternal Stress during the COVID-19 Pandemic and Infant Regulatory Capacity at 3 Months: A Longitudinal Study. Dev Psychopathol 2021, 1–9, doi:10.1017/S0954579421000766.

15. Manning, K.Y.; Long, X.; Watts, D.; Tomfohr-Madsen, L.; Giesbrecht, G.F.; Lebel, C. Prenatal Maternal Distress During the COVID-19 Pandemic and Associations With Infant Brain Connectivity. Biol Psychiatry 2022, doi:10.1016/J.BIOPSYCH.2022.05.011.

16. Lu, Y.-C.; Andescavage, N.; Wu, Y.; Kapse, K.; Andersen, N.R.; Quistorff, J.; Saeed, H.; Lopez, C.; Henderson, D.; Barnett, S.D.; et al. Maternal Psychological Distress during the COVID-19 Pandemic and Structural Changes of the Human Fetal Brain Plain Language Summary., doi:10.1038/s43856-022-00111-w.

17. Varescon, I.; Leignel, S.; Poulain, X.; Gerard, C. Coping Strategies and Perceived Stress in Pregnant Smokers Seeking Help for Cessation. J Smok Cessat 2011, 6, 126–132, doi:10.1375/JSC.6.2.126.

18. Razurel, C.; Kaiser, B.; Sellenet, C.; Epiney, M. Relation between Perceived Stress, Social Support, and Coping Strategies and Maternal Well-Being: A Review of the Literature. Women Health 2013, 53, 74–99, doi:10.1080/03630242.2012.732681.

19. Rimal, S.P.; Thapa, K.; Shrestha, R. Psychological Distress and Coping among Pregnant Women during the COVID 19 Pandemic. J Nepal Health Res Counc 2022, 20, 234–240, doi:10.33314/JNHRC.V20I01.4063.

20. Kinser, P.A.; Jallo, N.; Amstadter, A.B.; Thacker, L.R.; Jones, E.; Moyer, S.; Rider, A.; Karjane, N.; Salisbury, A.L. Depression, Anxiety, Resilience, and Coping: The Experience of Pregnant and New Mothers during the First Few Months of the COVID-19 Pandemic. J Womens Health 2021, 30, 654–664, doi:10.1089/JWH.2020.8866/ASSET/IMAGES/LARGE/JWH.2020.8866_FIGURE1.JPEG.

21. Werchan, D.M.; Hendrix, C.L.; Ablow, J.C.; Amstadter, A.B.; Austin, A.C.; Babineau, V.; Anne Bogat, G.; Cioffredi, L.A.; Conradt, E.; Crowell, S.E.; et al. Behavioral Coping Phenotypes and Associated Psychosocial Outcomes of Pregnant and Postpartum Women during the COVID-19 Pandemic. Scientific Reports 2022 12:1 2022, 12, 1–12, doi:10.1038/s41598-022-05299-4.

22. Thomason, M.; Graham, A.; Sullivan, E.; Heuvel, M.I. van den COVID-19 and Perinatal Experiences Study Available online: https://osf.io/uqhcv/ (accessed on 28 August 2022).

23. Brief Symptom Inventory - PsycNET Available online: https://psycnet.apa.org/doiLanding?doi=10.1037%2Ft00789-000 (accessed on 16 September 2022).

24. Blevins, C.A.; Weathers, F.W.; Davis, M.T.; Witte, T.K.; Domino, J.L. The Posttraumatic Stress Disorder Checklist for DSM-5 (PCL-5): Development and Initial Psychometric Evaluation. J Trauma Stress 2015, 28, 489–498, doi:10.1002/JTS.22059.

25. Carver, C.S. You Want to Measure Coping but Your Protocol’s Too Long: Consider the Brief COPE. Int J Behav Med 1997, 4, 92–100, doi:10.1207/S15327558IJBM0401_6.

26. Carver, C.S.; Scheier, M.F.; Weintraub, J.K. Assessing Coping Strategies: A Theoretically Based Approach. J Pers Soc Psychol 1989, 56, 267–283, doi:10.1037//0022-3514.56.2.267.

27. Connor-Smith, J.K.; Flachsbart, C. Relations between Personality and Coping: A Meta-Analysis. J Pers Soc Psychol 2007, 93, 1080–1107, doi:10.1037/0022-3514.93.6.1080.

28. García, F.E.; Barraza-Peña, C.G.; Wlodarczyk, A.; Alvear-Carrasco, M.; Reyes-Reyes, A. Psychometric Properties of the Brief-COPE for the Evaluation of Coping Strategies in the Chilean Population. Psicologia: Reflexao e Critica 2018, 31, 1–11, doi:10.1186/S41155-018-0102-3/TABLES/7.

29. Carver, C.S.; Scheier, M.F.; Weintraub, J.K. Assessing Coping Strategies: A Theoretically Based Approach. J Pers Soc Psychol 1989, 56, 267–283, doi:10.1037//0022-3514.56.2.267.

30. Lefmann, T.; Combs-Orme, T. Prenatal Stress, Poverty, and Child Outcomes. Child and Adolescent Social Work Journal 2014 31:6 2014, 31, 577–590, doi:10.1007/S10560-014-0340-X.

31. Silverman, M.E.; Medeiros, C.; Burgos, L. Early Pregnancy Mood before and during COVID-19 Community Re-strictions among Women of Low Socioeconomic Status in New York City: A Preliminary Study. Arch Womens Ment Health 2020, 23, 779, doi:10.1007/S00737-020-01061-9.

32. Sandel, M.; Faugno, E.; Mingo, A.; Cannon, J.; Byrd, K.; Garcia, D.A.; Collier, S.; McClure, E.; Jarrett, R.B. Neigh-borhood-Level Interventions to Improve Childhood Opportunity and Lift Children Out of Poverty. Acad Pediatr 2016, 16, S128–S135, doi:10.1016/J.ACAP.2016.01.013.

33. Acevedo-Garcia, D.; McArdle, N.; Hardy, E.F.; Crisan, U.I.; Romano, B.; Norris, D.; Baek, M.; Reece, J. The Child Opportunity Index: Improving Collaboration between Community Development and Public Health. Health Aff 2014, 33, 1948–1957, doi:10.1377/HLTHAFF.2014.0679/ASSET/IMAGES/LARGE/2014.0679FIGEX5.JPEG.

34. Ebner, M.; Wang, G.; Li, W.; Aertsen, M.; Patel, P.A.; Aughwane, R.; Melbourne, A.; Doel, T.; Dymarkowski, S.; de Coppi, P.; et al. An Automated Framework for Localization, Segmentation and Super-Resolution Reconstruction of Fetal Brain MRI. Neuroimage 2020, 206, 116324, doi:10.1016/J.NEUROIMAGE.2019.116324.

35. Gholipour, A.; Rollins, C.K.; Velasco-Annis, C.; Ouaalam, A.; Akhondi-Asl, A.; Afacan, O.; Ortinau, C.M.; Clancy, S.; Limperopoulos, C.; Yang, E.; et al. A Normative Spatiotemporal MRI Atlas of the Fetal Brain for Automatic Segmentation and Analysis of Early Brain Growth. Scientific Reports 2017 7:1 2017, 7, 1–13, doi:10.1038/s41598-017-00525-w.

36. Avants, B.B.; Tustison, N.J.; Song, G.; Cook, P.A.; Klein, A.; Gee, J.C. A Reproducible Evaluation of ANTs Similari-ty Metric Performance in Brain Image Registration. Neuroimage 2011, 54, 2033–2044, doi:10.1016/J.NEUROIMAGE.2010.09.025.

37. Smith, S.M. Fast Robust Automated Brain Extraction. Hum Brain Mapp 2002, 17, 143–155, doi:10.1002/HBM.10062.

38. Cox, R.W. AFNI: Software for Analysis and Visualization of Functional Magnetic Resonance Neuroimages. Computers and Biomedical Research 1996, 29, 162–173, doi:10.1006/cbmr.1996.0014.

39. Amin, S.B.; Orlando, M.S.; Dalzell, L.E.; Merle, K.S.; Guillet, R. Morphological Changes in Serial Auditory Brain Stem Responses in 24 to 32 Weeks’ Gestational Age Infants during the First Week of Life. Ear Hear 1999, 20, 410–418, doi:10.1097/00003446-199910000-00004.

40. Jiang, Z.D.; Xiu, X.; Brosi, D.M.; Shao, X.M.; Wilkinson, A.R. Sub-Optimal Function of the Auditory Brainstem in Term Infants with Transient Low Apgar Scores. Clinical Neurophysiology 2007, 5, 1088–1096, doi:10.1016/J.CLINPH.2007.01.018.

41. DiPietro, J.A.; Kivlighan, K.T.; Costigan, K.A.; Rubin, S.E.; Shiffler, D.E.; Henderson, J.L.; Pillion, J.P. Prenatal An-tecedents of Newborn Neurological Maturation. Child Dev 2010, 81, 115–130, doi:10.1111/J.1467-8624.2009.01384.X.

42. de Asis-Cruz, J.; Krishnamurthy, D.; Zhao, L.; Kapse, K.; Vezina, G.; Andescavage, N.; Quistorff, J.; Lopez, C.; Limperopoulos, C. Association of Prenatal Maternal Anxiety With Fetal Regional Brain Connectivity. JAMA Netw Open 2020, 3, e2022349–e2022349, doi:10.1001/JAMANETWORKOPEN.2020.22349.

43. Garrett, D.D.; Kovacevic, N.; McIntosh, A.R.; Grady, C.L. The Importance of Being Variable. J Neurosci 2011, 31, 4496–4503, doi:10.1523/JNEUROSCI.5641-10.2011.

44. Rogers, C.E.; Lean, R.E.; Wheelock, M.D.; Smyser, C.D. Aberrant Structural and Functional Connectivity and Neurodevelopmental Impairment in Preterm Children. J Neurodev Disord 2018, 10, doi:10.1186/S11689-018-9253-X.

45. Evans, T.M.; Kochalka, J.; Ngoon, T.J.; Wu, S.S.; Qin, S.; Battista, C.; Menon, V. Brain Structural Integrity and In-trinsic Functional Connectivity Forecast 6 Year Longitudinal Growth in Children’s Numerical Abilities. J Neuro-sci 2015, 35, 11743–11750, doi:10.1523/JNEUROSCI.0216-15.2015.

46. Barch, D.M.; Belden, A.C.; Tillman, R.; Whalen, D.; Luby, J.L. Early Childhood Adverse Experiences, Inferior Frontal Gyrus Connectivity, and the Trajectory of Externalizing Psychopathology. J Am Acad Child Adolesc Psychiatry 2018, 57, 183–190, doi:10.1016/J.JAAC.2017.12.011.

47. Thomason, M.E.; Hect, J.L.; Waller, R.; Curtin, P. Interactive Relations between Maternal Prenatal Stress, Fetal Brain Connectivity, and Gestational Age at Delivery. Neuropsychopharmacology 2021 46:10 2021, 46, 1839–1847, doi:10.1038/s41386-021-01066-7.

48. Han, L.; Bai, H.; Lun, B.; Li, Y.; Wang, Y.; Ni, Q. The Prevalence of Fear of Childbirth and Its Association With In-tolerance of Uncertainty and Coping Styles Among Pregnant Chinese Women During the COVID-19 Pandemic. Front Psychiatry 2022, 13, doi:10.3389/FPSYT.2022.935760.

49. Anderson, M.R.; Salisbury, A.L.; Uebelacker, L.A.; Abrantes, A.M.; Battle, C.L. Stress, Coping and Silver Linings: How Depressed Perinatal Women Experienced the COVID-19 Pandemic. J Affect Disord 2022, 298, 329–336, doi:10.1016/J.JAD.2021.10.116.

50. Giurgescu, C.; Penckofer, S.; Maurer, M.C.; Bryant, F.B. Impact of Uncertainty, Social Support, and Prenatal Cop-ing on the Psychological Well-Being of High-Risk Pregnant Women. Nurs Res 2006, 55, 356–365, doi:10.1097/00006199-200609000-00008.

