## Supplemental Table 1 for "Impact of COVID-19 related maternal stress on fetal brain development: A Multimodal MRI study"

| Default Atlas Labels | Mapped Brain Structure |
| --- | --- |
| Midbrain_L | Brainstem |
| Cerebellum_L, Cerebellum_R | Cerebellum |
| Cortical_Plate_L, Cortical_Plate_R | Cortical Plate |
| Caudate_L, Caudate_R, Putamen_R, Putamen_L, Thalamus_L, Thalamus_R,  Fornix, Internal_Capsule_L, Internal_Capsule_R, Subthalamic_Nuc_L,  Subthalamic_Nuc_R, Hippocampal_Comm | Deep Grey |
| CSF | Extra Axial CSF |
| Amygdala_L, Amygdala_R, Hippocampus_L, Hippocampus_R | Hippocampus, Amygdala Complex |
| Lateral_Ventricle_L, Lateral_Ventricle_R | Intra-ventricular CSF |
| White_Matter_L, White_Matter_R, Inter_Zone_L, Inter_Zone_R,  Subplate_L, Subplate_R, Vent_Zone_L, Vent_Zone_R, Corpus Callosum | White Matter |
| All Structures Listed Above | Total Brain Volume |

Supplemental Table 1. Mappings used from the original Gholipour atlas to the structures studied during this analysis.
